# RT-qPCR-based tests for SARS-CoV-2 detection in pooled saliva samples for massive population screening to monitor epidemics

**DOI:** 10.1101/2021.05.06.21256733

**Authors:** Michał Różański, Aurelia Walczak-Drzewiecka, Jolanta Witaszewska, Ewelina Wójcik, Arkadiusz Guziński, Bogumił Zimoń, Rafał Matusiak, Joanna Kazimierczak, Maciej Borowiec, Katarzyna Kania, Edyta Paradowska, Jakub Pawełczyk, Jarosław Dziadek, Jarosław Dastych

## Abstract

Swab, quantitative, reverse transcription polymerase chain reaction (RT-qPCR) tests remain the gold standard of diagnostics of SARS-CoV-2 infections. However, these tests are costly and time-consuming, and swabbing limits their throughput. We developed a 3-gene, seminested RT-qPCR test with SYBR green-based detection, optimized for testing pooled saliva samples for high-throughput diagnostics of epidemic-affected populations. The proposed two-tier approach depends on decentralized self-collection of saliva samples, pooling, 1^st^-tier testing with the mentioned highly sensitive screening test and subsequent 2^nd^-tier testing of individual samples from positive pools with the *in vitro* diagnostic (IVD) test. The screening test was able to detect 5 copies of the viral genome in 10 µl of isolated RNA with 50% probability and 18.8 copies with 95% probability and reached Ct values that were highly linearly RNA concentration-dependent. In the side-by-side comparison (testing artificial pooled samples), the screening test attained slightly better results than the commercially available IVD-certified RT-qPCR diagnostic test (100% specificity and 89.8% sensitivity vs. 100% and 73.5%, respectively). Testing of 1475 individual clinical samples pooled in 374 pools of 4 revealed 0.8% false positive pools and no false negative pools. In weekly prophylactic testing of 113 people within 6 months, a two-tier testing approach enabled the detection of 18 infected individuals, including several asymptomatic individuals, with a fraction of the costs of individual RT-PCR testing.

## Introduction

In the spring of 2020, the world faced the emerging threat of the novel coronavirus SARS-CoV-2 causing COVID-19 disease. In a relatively short time since its identification, this virus led to a COVID-19 pandemic that, according to the WHO Report of March 02, 2021, reached over 140 million cases and 3 million deaths (https://www.who.int/publications/m/item/weekly-epidemiological-update-on-covid-19---20-april-2021, Access April 20, 2021). SARS-CoV-2 belongs to the β-coronavirus genus, and its genetic material is positive-sense single-stranded RNA of nearly 30 kilobases in length. Such a global pandemic is unprecedented in the age of molecular biology and molecular test-based epidemiology. This has resulted in the worldwide research and development of new approaches for diagnostics, vaccination and treatment to control rapidly evolving epidemic situations. In March 2020, the first vaccines, including those based on mRNA and genetically modified adenoviruses, were approved for use and introduced in population-wide vaccination programs^1,2^. Until then, diagnostic tests along with social distancing biosafety protection were the only methods to control the epidemic. The first employed diagnostic method for SARS-CoV-2 was RT-qPCR-based identification of the virus’s genetic material in nasopharyngeal swabs, and RT-PCR-based virus detection is still considered the gold standard in COVID-19 testing^3^. While at least 1.9 billion tests have been performed worldwide (Retrieved from: https://www.worldometers.info/coronavirus/ access April 04, 2021), it was perhaps not enough to significantly limit the global spread of SARS-CoV-2.However,this massive diagnostic effort provided critical data supporting public health efforts. There are examples showing the possibility of locally reducing the scale of the epidemic by combining mass testing for the virus with isolating infected people and taking other safety measures. For example, an analysis of testing-based public health activities in Wuhan shows that the spread of the virus has slowed down in the population. The time of doubling the number of patients first increased from 2 to 4 days and then even to 19 days^4^. Similarly results were observed in South Korea. The adopted model of fighting the epidemic contributed to gaining control over the coronavirus in just 3 weeks thanks to, inter alia, mass testing of the population^5^. The introduction of mass testing in Australia made it possible to keep the incidence low and even eliminate new cases (https://www.health.gov.au/news/health-alerts/novel-coronavirus-2019-ncov-health-alert/coronavirus-covid-19-current-situation-and-case-numbers access April 04, 2021), similar to Israel^6^. Thus, one potentially effective strategy of controlling SARS-CoV-2 spreading by anti-epidemic measures other than vaccination is mass-scale testing that allows as many people as possible to be tested as quickly as possible.

We decided to develop a new screening RT-qPCR-based test dedicated to the massive screening needed to monitor spreading infections in populations during pandemic crises. The specific goals were a) high sensitivity; b) suitability for quantitative measurements; c) good performance in biologic samples pooled from multiple individuals d) open-source (at minimal level competes for unique resources with other diagnostics) adaptable to different locally available sources of test components. The last goal reflected our experience with a pandemic-related impact on the availability of dedicated commercial diagnostic tests. As a result, we developed a seminested RT-qPCR SARS-CoV-2 test based on SYBR green detection that meets the above criteria. Furthermore, we have developed a protocol for routine repetitive testing of large groups of individuals allowing us to perform such tasks at a significantly lower cost than regular PCR-based diagnostic testing. This protocol employs saliva, rather than swabs, as self-collected saliva samples are well suited for massive pooling tests.

## Materials and Methods

### Biological material

The usage of anonymized samples from patients of the Center of Molecular Diagnostics of Pathogens, Proteon Pharmaceuticals S.A. in the study was approved by the Bioethical Committee at the University of Lodz (Res. nr 3/KBBN-UŁ/I/2020-21). The study was conducted in accordance with the Declaration of Helsinki and the good clinical practice guidelines. Written informed consent was obtained from all participants before study entry. Titrated, heat-inactivated SARS-CoV-2 virus was obtained from BEI Resources, Catalog No. NR-52286. Samples of genomic RNA isolated from human coronaviruses HCoV 229E and HCoV NL63 and respiratory syncytial viruses RSV A2001/3-12 and RSV B1 were obtained from BEI Resources. Titrated SARS-CoV-2 RNA isolated from Vero cell culture was a kind gift from Prof. Krzysztof Pyrć (Jagiellonian University).

### Reagents

SuperScript IV Reverse Transcriptase (CN: 18090200), dNTP mix 10 mM (CN: R0193), RNaseOUT™ Recombinant Ribonuclease Inhibitor (CN: 10777019) and DEPC-treated water (CN: 750024) were purchased from Thermo Fisher Scientific. DNA Taq Polymerase (CN: E2500) and attached 10x Polymerase Buffer B (with 15 mM MgCl_2_) were purchased from EURx. LightCycler® 480 SYBR Green I Master mix (CN: 04887352001) was purchased from Roche, and the Maxwell® RSC Viral Total Nucleic Acid Purification Kit (CN: AS1330) was purchased from Promega.

DiaPlexQ™ Novel Coronavirus (2019-nCoV) Detection Kit (CN: SQD52-K100; SolGent Co. Ltd.) and 2019-Novel Coronavirus (2019-nCoV) Triplex RT-qPCR Detection Kit (CN: CD302-02; Vazyme) were used as reference IVD tests for SARS-CoV-2 detection.

### Sample collection, pool design and RNA isolation

Saliva samples in 5 ml polypropylene tubes packed in double plastic bags were first heat inactivated at 90°C for 15 min in a dry oven. Following inactivation, pooling samples were created by combining four 75 μl aliquots of individual saliva samples into one 300 μl sample, and either pooled or individual samples were processed according to the instructions of isolation RNA using the Maxwell® RSC Viral Total Nucleic Acid Purification Kit with the Maxwell® RSC 48 Instrument (Promega). Briefly, 300 μl saliva was mixed with 330 µl lysis buffer containing proteinase K, incubated at 56°C for 10 min and briefly centrifuged. Next, each sample was applied to separate wells of disposable cartridges in an RNA Maxwell® RSC 48 Instrument for semi-automatic RNA extraction.

### Choice of conservative regions in SARS-CoV-2 genome

First, 94155 genome sequences were downloaded from the GisAid database, including 94139 SARS-CoV-2 and 16 non-SARS-CoV-2 genomes (Appendix 1, access 07.09.2020). Then, one reference genome (NC_045512.2) was chosen from which sequences of RdRp, Spike and N genes were selected. Using BLAST 2.9.0+ (default parameters) and reference gene sequences as queries, desired genes were extracted from the remaining SARS-CoV-2 genomes. Sequences were aligned in MAFFT(v7.310) with adjustment of direction and the FFT-NS-1 method to build a full MSA (multiple sequence alignment) and to find conservative regions among selected sequences. Moreover, as a control for the reaction, consensus sequences of the human GAPDH gene were extracted from 5 transcriptional variants: 1,2,3,4 and 7 (NM_002046.7, NM_001256799.3, NM_001289745.3, NM_001289746.2 and NM_001357943.2).

### Primer design and verification

Sets of primers (Table 1) targeting the mentioned fragments were designed using the open access software UGENE (v.35). Then, the primers’ sensitivity and specificity were verified. For this purpose, all *Coronoviridae* genomes marked as complete were downloaded from the nucleotide database (NCBI). First, non-SARS-CoV-2 genomes were chosen for specificity analysis, resulting in 2802 sequences. Then, from all SARS-CoV-2 genomes, any incomplete duplicates or sequences shorter than 90% of the reference NC_045512.2 genome were deleted, and 80770 SARS-CoV-2 were used for the sensitivity analysis. *In silico* PCR was conducted (https://github.com/egonozer/in_silico_pcr, accessed November 2020) with the mismatch threshold set to1 bp. In this way, a set of primers (both seminested and detection primers) was chosen. Primers were synthesized by Genomed, Poland and purified by high-performance liquid chromatography. Parameters of primers (GC content, Tm and self-complementarity) were analyzed in Oligo Calc (http://biotools.nubic.northwestern.edu/OligoCalc.html access September 2020). Primers used in the screening test were routinely premixed and stored as ready-to-use stock solutions with each primer concentration of 10 µM in ultrapure water; details are listed in Table 2.

**Table 1.**
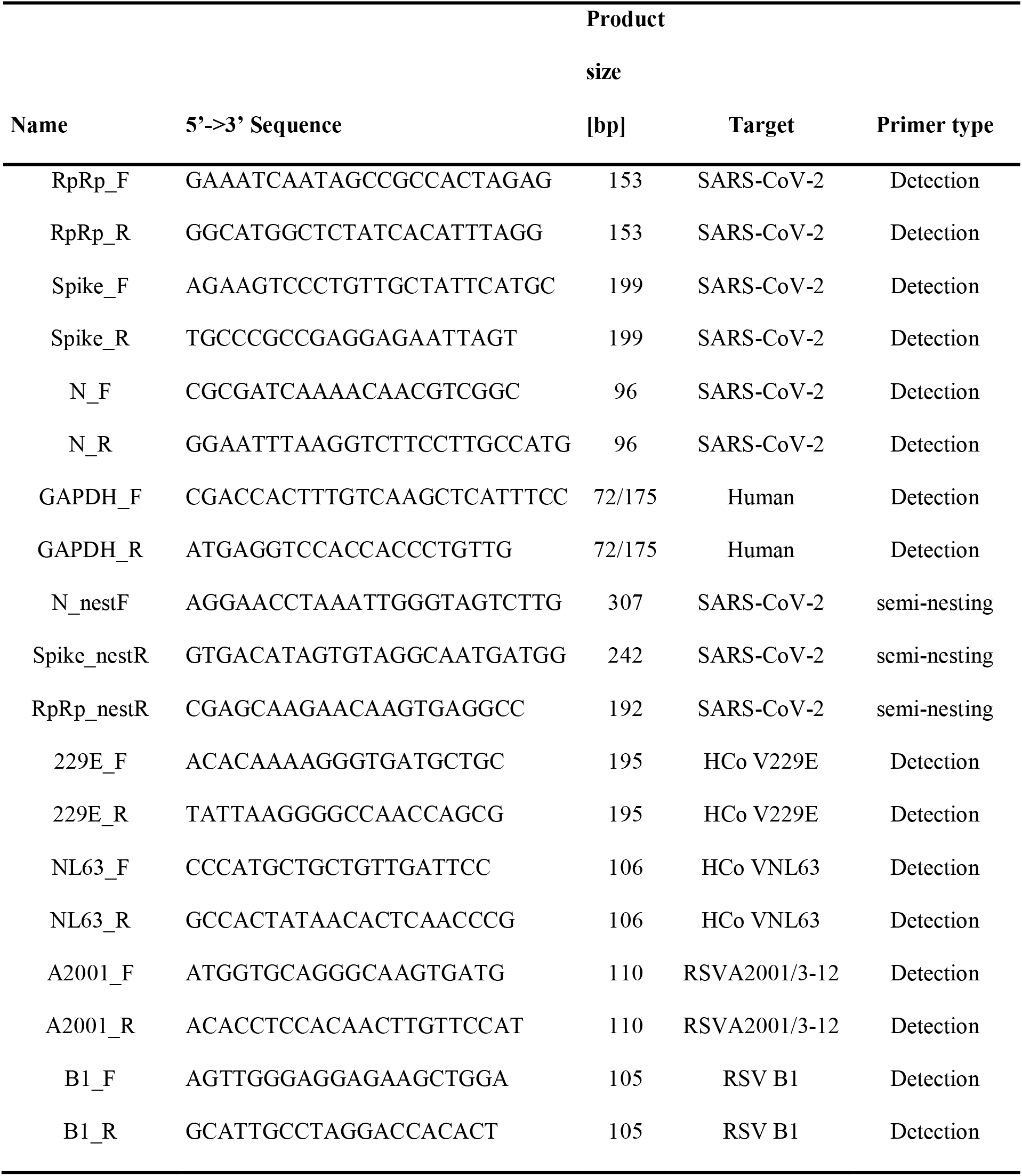
Primers used in the study

**Table 2.** Primer mixes used in this study

| Primer mix name | Content |
| --- | --- |
| Primer mix 1 | N_R, Spike_nestR, RdRp_nestR, GAPDH_R |
| Primer mix 2 | N_nestF, Spike_F, RdRp_F |
| GAPDH mix | GAPDH_F, GAPDH_R |
| RdRp mix | RdRp_F, RdRp_R |
| Spike mix | Spike_F, Spike_R |
| N mix | genN_F, genN_R |

### Design of seminested RT-PCR

The first step of the procedure consisted of reverse transcription of viral RNA and 20 cycles of multiplex PCR. First, 5 µl of Mastermix 1 (Table 3) and 10 µl of RNA solution were added to each well in a 96-well PCR plate. The plate was sealed, incubated in a thermal cycler for 4 minutes at 65°C (initial denaturation of RNA) and 1 minute at 56°C (reverse primers annealing), and then immediately chilled to 4°C in a cooling block. Next, the sealing film was removed, and 10 µl of Mastermix 2 (Table 3) was added to each well. The plate was sealed again, and thermal cycling was performed, as presented in Table 4. The PCR product was diluted 30x in ultrapure water in a standard flat-bottomed 96-well plate, shaken for 5 minutes at 750 rpm on an orbital plate shaker and used in the second nested qPCR reaction.

**Table 3.**
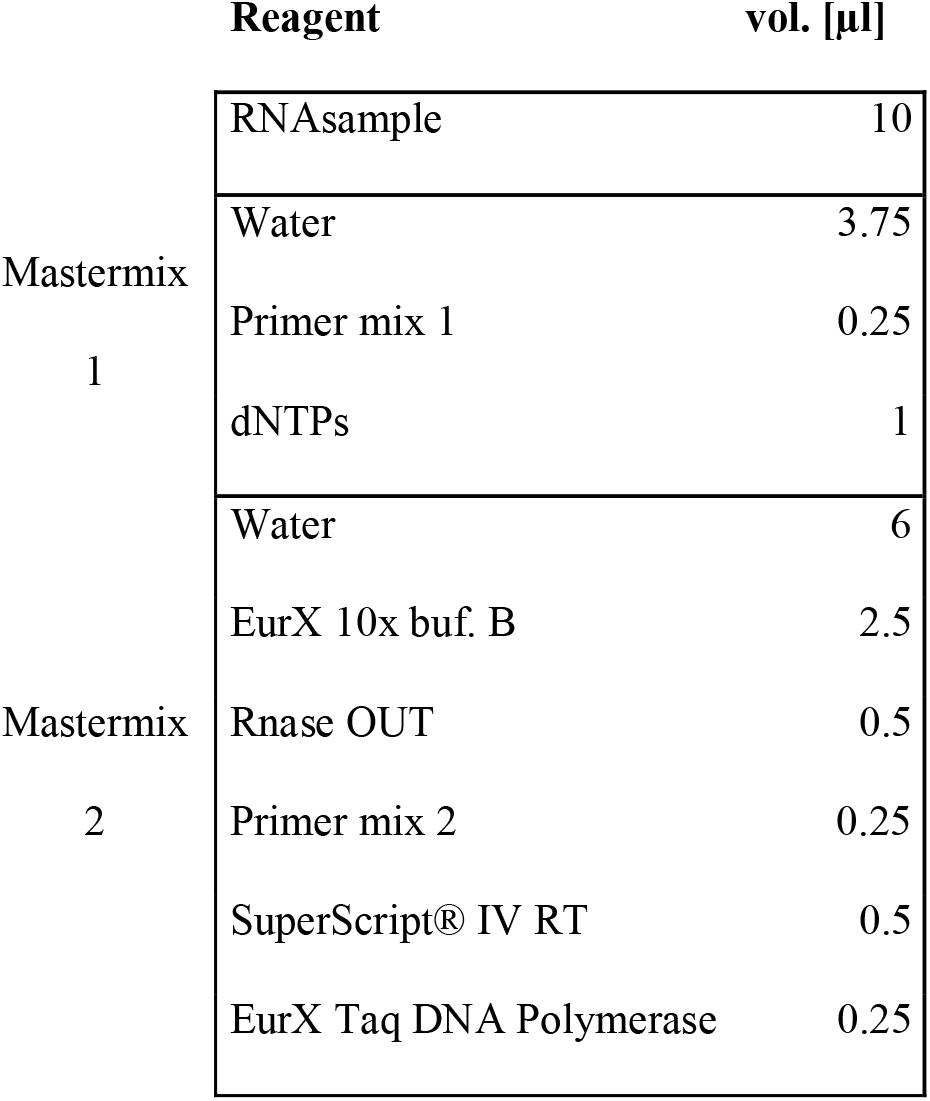
Composition of the reaction mix in the first step of the reaction

**Table 4.**
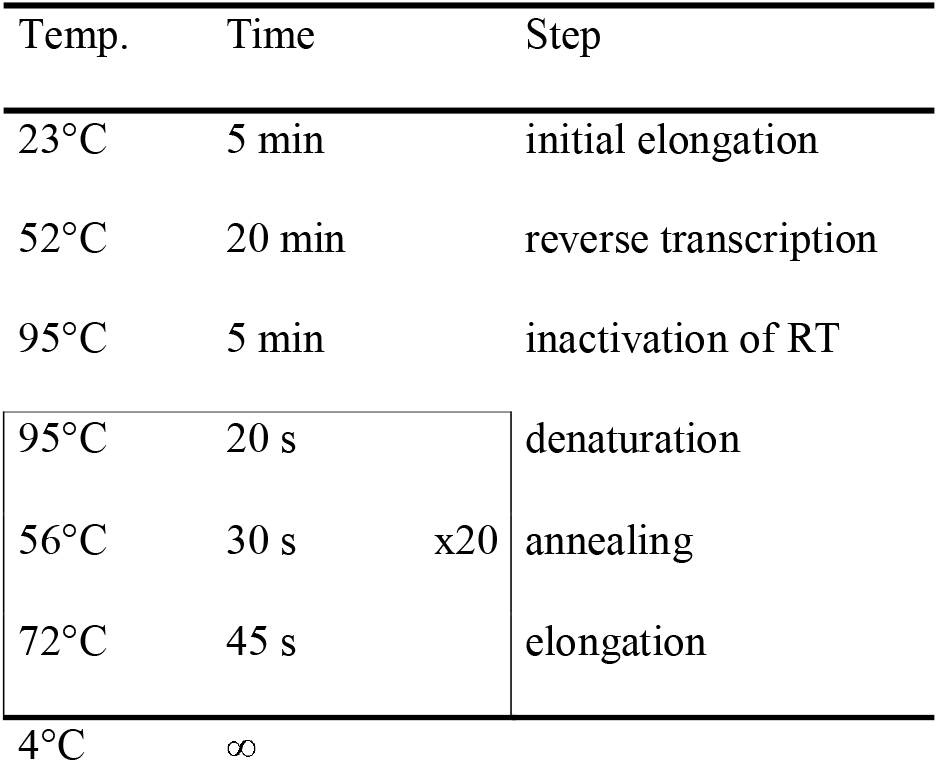
Thermal profile of the first PCR

Nested qPCR of individual viral genes and human controls was performed in duplicate in separate wells of 384-well plates. Each reaction mix consisted of 5 µl of LightCycler® 480 SYBR Green I Master Mix (2x), 0.3 µl of proper detection primer mix (Table 2), 1.7 µl of water and 3 µl of 30x diluted product of the first PCR. Detection plates were assembled by an automatic pipetting station (FasTrans, Analytik Jena AG). qPCR was performed on a LightCycler® 480 Instrument II (Roche) with the following cycling conditions: 5 min of initial denaturation at 95°C; 45 cycles of 10 s at 95°C, 12 s at 56°C and 20s at 72°C with SYBR green fluorescence measurement; product melting curve measurement was performed with 8 measurements per °C.

### Cross reactivity with RNA of selected respiratory viruses

Genomic RNA standards of human respiratory viruses were used according to the manufacturer’s instructions. 5 µl of HCoV 229E, 10 µl HCoV NL63, 2 µl RSV A2001/3-12, and 5 µl RSV B1 were added per reaction and tested according to the main protocol of the method. As a positive control, additional reactions were performed with primers specific for these viruses (Table 1). In control reactions, RNA was transcribed to cDNA with a single reverse primer. The first PCR step was omitted, and 3 µl of undiluted cDNA sample was directly used as a matrix for qPCR.

### RNA-based Ct calibration curve

Titrated SARS-CoV-2 RNA isolated from infected Vero cell culture was serially diluted in ultrapure water. At each concentration, reverse transcription coupled with seminested PCR was performed in duplicate, followed by qPCR in triplicate for each gene, resulting in 18 qPCR replicates in total and 6 for each individual gene. Only data points with correct melting temperatures were employed for further analysis. Ct values were plotted against logarithmic concentration of viral RNA, and linear regression curves were fitted to data using GraphPad Prism software.

### Establishing criteria for SARS-CoV-2 detection in screening tests

First, melting curves from both reference viral RNA standards and from clinical samples with high probability of being true positives were used to determine the melting temperature range for specific PCR products in qPCR outcome for viral gene detection. Next, the Ct threshold distinguishing positive from negative qPCR outcomes for viral gene detection was determined by parallel analysis of RNA from a set of pooled saliva samples with a screening test and reference test (DiaPlexQ™). To this end, 7 representative SARS-CoV-2-positive saliva samples (with different viral loads) and 20 negative saliva samples were used. Each positive sample was pooled by combining 3, 5 or 7 randomly chosen negative samples. Each pool was prepared in duplicate (but with different negative samples). All prepared pools as well as individual saliva samples (positive and negative) were isolated according to the described protocol and tested with the screening and reference test. Data from this experiment were also employed to determine the number of positive qPCR replicates needed to consider the clinical pooled saliva sample as positive.

### Estimation of the probability of obtaining a positive result

Data points from calibration curve preparation were extracted. For each tested concentration of viral RNA, the number of positive (determined according to criteria of SARS-CoV-2 detection in screening test) replicates of viral genes (N, spike and RdRp) was divided by the total number of tested viral replicates (n=18). The result is an observed probability 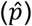 of obtaining a positive replicate of the viral gene at a given concentration of viral RNA. As the criteria of positive screening tests may assign to each replicate only a value of 1 or 0, we assumed that the variable was Bernoulli’s binomial distribution. We used the normal distribution approximation approach to calculate the 95% confidence interval (CI) of the data:

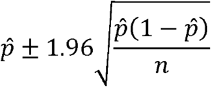

where *p* is an observed probability, 1.96 is a coefficient for the 95% confidence interval and n is the number of tested samples. Subsequently, values for upper and lower CI for each point were recalculated into probability of obtaining positive result of whole test (probability of obtaining at least 4 positive replicates out of total 6 replicates at given *p*):

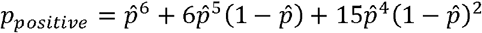

whereis 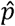 is an observed probability for a single replicate and p_positive_ is the probability for the whole test to give a positive result. Newly calculated intervals were plotted against logarithmic concentration of viral RNA, and a four-parameter logistic curve was used to calculate P_50_ and P_95_ (concentration at which test has 50% and 95% chance to give positive outcome, respectively). Curve fitting was performed in GraphPad Prism software.

### RNA isolation efficiency

From a group of SARS-CoV-2 negative (confirmed by 2019-Novel Coronavirus (2019-nCoV) Triplex RT-qPCR diagnostic test, Vazyme), heat inactivated, frozen saliva samples, 21 specimens were randomly chosen. Subsequently, samples were divided into seven three-element groups, and saliva within groups was equally pooled and mixed, forming 7 averaged saliva representations. Each pooled sample was aliquoted by 500 µl into 4 test tubes, and 10 µl of HBSS (control) or proper dilution of standard: titrated, heat-inactivated SARS-CoV-2 virus in HBSS was added. Next, RNA was isolated, and a screening test was performed. RNA isolation efficiency was calculated based on measured Ct values and the Ct calibration curve; theoretical viral load was calculated for each gene replicate. Subsequently, that value was multiplied by the volume coefficient of 26.7 (which converts [n/10 µl of RNA] into [n/ml of saliva]: 10 µl of RNA sample out of 80 µl of RNA isolate from 300 µl of saliva sample) and divided by the initial viral load in the saliva sample.

### Economic analysis of pooled screening test

Center of Molecular Diagnostics of Pathogens, Proteon Pharmaceuticals S.A. provided information on costs included in testing viral samples according to the gold standard diagnostic approach (based on commercially available one-step RT-qPCR diagnostic kits, isolation of RNA, disposables, PPE, labor and other direct costs of diagnostics). The summarized costs were considered as reference 100% - the whole cost of standard diagnostic test per sample. Subsequently, the cost of the screening test was calculated based on the following premises: 1) positive cases were randomly distributed among the tested population with an even distribution; 2) the test had ideal sensitivity and specificity; and 3) if the pool provided a positive result, each sample within the pool was subsequently tested individually with reference, validated, diagnostic tests. Three pooling factors were chosen for analysis: 4-, 8- and 12-fold. For each case, the baseline cost of the screening test is the cost of testing a pool with a screening test (seminested qPCR with SYBR green detection) divided by a pooling factor. Then, the number of positive pools in the tested population is assessed based on the percent of positive individuals in the population and the chosen pooling factor (with the use of elementary combinatorics equations). The number of positive pools was next multiplied by the pooling factor and cost of the diagnostic test per sample, and the result was divided by the number of individual samples in the tested population. Such a variable component was added to a baseline, and the result was the total cost of the screening test.

## Results

### Validation of primer design

Bioinformatic specificity analysis revealed that the chosen detection primers were specific to 99.857% of the analyzed *Coronaviridae* genomes with four false positive results, while this test was 99.995% sensitive towards SARS-CoV-2 genomes with four negative results. A summary of this information is presented in the confusion matrix in Figure 1.

**Figure 1.**
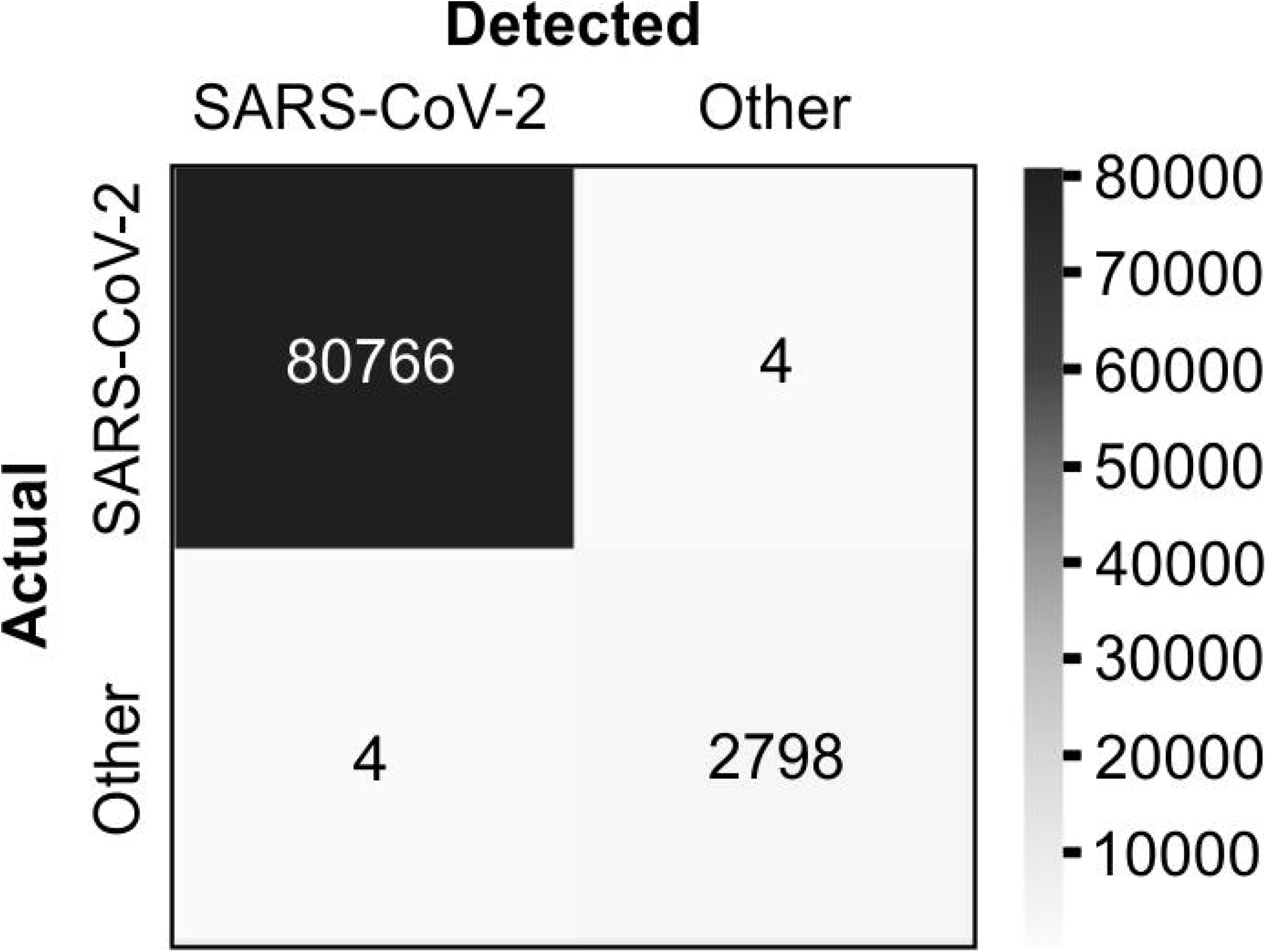
Specificity of primer pairs used in screening test. Bioinformatic confusion matrix of chosen primer pair sequences vs. SARS-CoV-2 genomes and other *Coronaviridae* genomes.

### Criteria of SARS-CoV-2 detection in screening test

Melting temperatures of specific qPCR products for viral genes: data from several independent experiments using several different matrix RNA sources (tittered standard RNA or RNA isolated from BEI standard or RNA isolated from clinical samples) were analyzed. Data from samples with high probability of being true positive measurements (both replicates similar, Tm close to preliminary measured values and Ct<30 cycle) were extracted. Melting temperatures for individual genes were as follows (mean ± 3xSD (°C), number of data points): RdRp 78.25±0.311, 140; spike 80.27±0.319, 140; N 80.72±0.322, 140; GAPDH 78.99±0.445, 344. Considering the screening purpose of the test, we intentionally chosen slightly wider threshold values to keep overall output oversensitive rather than overspecific, especially because weaker signals (higher Ct values) tended to deviate more. Finally, the chosen qualification rules for each gene were selected as follows: “to be considered specific, the measured melting temperature must lie within the following intervals (center of an interval ± deviation (°C)): 78.3±0.35 for RdRp; 80.3±0.35 for Spike; 80.7±0.35 for N; 79.0±0.50 for GAPDH”. Representative melting curves are presented in Figure 2A. The thermal cycler used in this study (LightCycler 480) always saves a colder peak as Tm1 and a warmer peak as Tm2 in the case of a two-product melting curve, regardless of the height of such peaks; if the case of two products occurred, we checked both measured melting temperatures against the chosen threshold. In the next series of experiments, seven SARS-CoV-2-positive saliva samples were pooled in duplicate with a pooling factor of 4-, 6- or 8-fold, with randomly chosen negative samples (as described in Materials and Methods), and RNA was isolated. The screening test was able to correctly identify 20/20 negative samples, 7/7 not pooled positive samples, 13/14 four-fold pooled positive samples, 12/14 six-fold pooled samples and 12/14 eight-fold pooled samples, which gives 100% specificity and 89.8% sensitivity within the experiment. As a reference, the validated diagnostic test DiaPlexQ^™^was able to identify 20/20 negative samples, 5/7 not pooled positives, and 10/14, 10/14 and 9/14 pooled positives, reaching 100% specificity and 73.5% sensitivity, respectively. To compare Ct values of each viral gene in pooled samples that were revealed as positive by screening test with Ct values in corresponding single positive samples, ΔCt values (number of cycles of delay in pooled samples) were calculated and presents as follows (pooling factor, mean±SD, n): P4, 3.28±2.73, 80; P6, 4.20±2.34, 72; P8, 4.17±1.81, 70. Data from this experiment were also used to optimize qualification criteria for the screening test to make it more sensitive or more selective (Figure 2B) by choosing the number of replicates of viral genes that need to be positive to consider a tested sample as positive. The optimal threshold rule was chosen to read as follows: “at least 4 of a total of 6 viral replicates must be positive (proper Tm and Ct values) to consider the tested sample as positive”.

**Figure 2.**
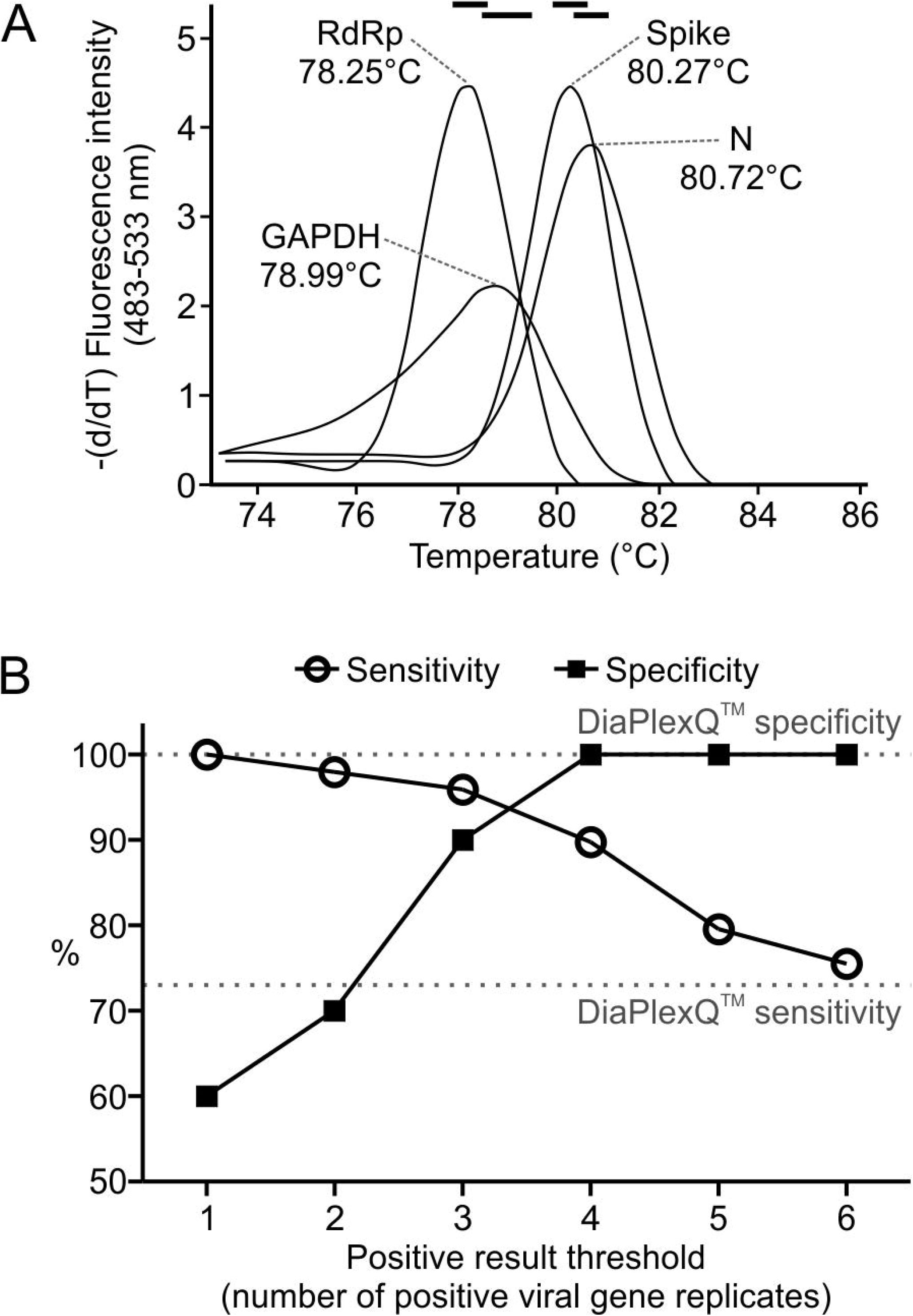
Criteria of the test result assignment. A: Derivatives of the representative melting curves of the tested genes in the screening test and mean Tm values. Black bars on top: temperature intervals within which the measured Tm value is considered specific for each gene. B: Sensitivity and specificity of the seminested SARS-CoV-2 screening test depending on the positive result criterion. The percentage of true positive samples (n=49) and true negative samples (n=20) which were qualified correctly depending on the number of positive replicates of viral genes (in total of 6) in the screening test that was chosen as a threshold. Dotted lines: respective parameters, achieved by validated diagnostic test.

### Quantitative performance of screening test

Tittered SARS-CoV-2 RNA was utilized to generate calibration curves for the Ct values of individual genes tested in the screening test. The results are presented in Figure 3A. For each gene across the whole tested concentration range, the relation between the obtained Ct values and logarithm of viral RNA concentration was highly linear. Slight difference between viral genes was observed. The RdRp product tended to result in somewhat smaller Ct values than the other two genes. This trend matches the results obtained in testing clinical saliva samples. After comparing 42 positive samples in which all 6 replicates of viral genes were positive, the Ct values were as follows (mean±SD, n=84): 21.67±5.66 for RdRp; 24.34±6.22 for Spike; 25.44±5.91 for N. The results for RdRp differed significantly (P<0.05) from those for the two other genes, between which there were no significant differences (ANOVA with Sheffé’s *post hoc* test).

**Figure 3.**
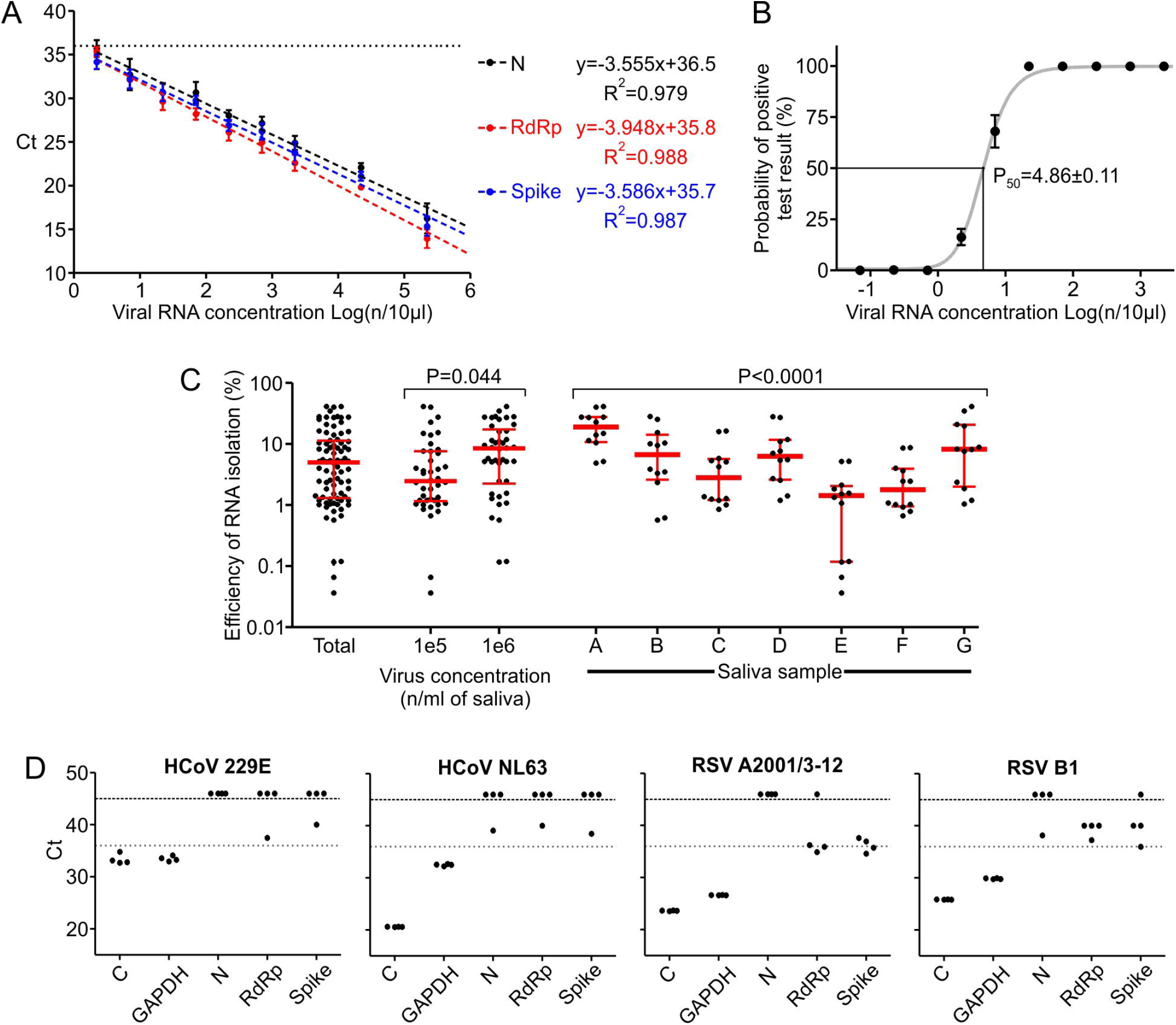
Quantitative performance of the screening test. A: RNA-based Ct calibration curves for individual genes. Mean±SD, n=6. Dashed lines: linear regression curves fitted to data. The calculated curve equations and coefficients of determination are listed on the right side. Linear fitting P<0.0001 for each gene. Dotted horizontal line: threshold Ct value for specific amplification. B: Probability of obtaining positive results of the screening test depending on viral RNA concentration in a sample. Black points/whiskers: 95% confidence intervals with the center of each interval marked. Gray line: four parameter logistic curve fitted to data. P_50_ value presented as the mean±SEM. C: Efficiency of RNA isolation: number of copies calculated from Ct calibration curve divided by number of viral particles added to saliva samples. Each point represents efficiency calculated independently from a single viral gene replicate. Red lines/whiskers: median and interquartile range. Data points presented as total are also analyzed by groups, depending on virus concentration or saliva sample used as a carrier for isolation. The significance of differences between groups was calculated with ANOVA. No *post hoc* tests were performed. D: Experimental verification of cross-reactivity with selected respiratory viruses. Ct values for individual genes obtained by qPCR in a seminested screening test using RNA samples of chosen non-SARS-CoV-2 respiratory viruses; C: control primer pair specific to each virus. Upper dashed line: last cycle of the reaction; points above indicate samples with no detectable amplification. Lower dotted line: chosen threshold for specific amplification; points above are considered nonspecific by default.

Based on Ct values for minimal RNA concentration that still gives detectable positive signal (from calibration curve experiment), a Ct value of 36 was chosen as a threshold value above which amplification was considered nonspecific by default. In the case of the control human gene (GAPDH) Ct values were extracted from several independent experiments in which RNA isolated from individual or pooled saliva samples was tested. GAPDH from 243 tested samples had a mean Ct value of 20.53 with an SD of 1.61, and the highest Ct value reached in the tested population was 26.39. Therefore, we decided to use a slightly higher threshold value of 28.0, above which, probably due to inefficiency of RNA isolation or too high of an inhibitor load in a sample, the result of the test is questionable.

Based on the results collected during the preparation of calibration curves, the probability of obtaining a positive result of the screening test, depending on the viral RNA concentration in the tested sample, was estimated. A logistic curve was fitted to the calculated probability values (Figure 3B); the P_50_ value was reached at a concentration of 4.86±0.11 genome copies per 10 µl of RNA sample (mean±SEM), and the P_95_ value was 18.8±0.75.

Next, we assessed the efficiency and reproducibility of viral RNA isolation. Seven representative saliva samples were used as carriers for 10^4^, 10^5^ and 10^6^ viral particles/ml; subsequently, RNA was isolated according to a standard protocol, and a screening test was performed. Only 54% of viral gene replicates at a concentration of 10^4^/ml were positive, and thus for further analysis, only two higher concentrations (which were 100% positive) were used. The results are presented in Figure 3C. The median RNA isolation efficiency was equal to 5.00%, with lower and upper quartiles equal to 1.33% and 11.09%, respectively. This value is somewhat dependent on the concentration of viral particles in the source material (3.42× higher median for the 10^6^/ml group than 10^5^/ml) and strongly dependent on individual saliva sample parameters (13.2× difference between the highest and lowest efficient saliva samples). To complete the methodology of quantitative measurements, an experimental cross reactivity test with RNA isolated from four respiratory viruses was performed and revealed no amplification in viral genes of the screening test. The results are presented in Figure 3D. None of the replicates of viral genes shown in a figure can be considered specific according to assumed test thresholds (Tm and Ct values). GAPDH, the control gene, showed specific amplification because the RNA samples were derived from human cell cultures.

### Economic analysis of pooled screening test

The results of the comparison of the cost of screening pooled test to individual (IVD certified) diagnostic testing are presented in Figure 4. This analysis suggests that pooling of samples has economic justification only when the predicted number of positive cases in the tested population is below 30%. At the higher prevalence of infection, cost of the pooling approach per sample exceeds the cost of the individual diagnostic tests and negatively affects workflow organization (additional RNA isolation and detection steps), significantly delaying the results. However, in the case of lower prevalence, the presented approach may significantly decrease the cost of testing large groups of people. Comparing different pooling factors, the 4-fold option seems to be the most cost effective, assuring the lowest overall cost in the widest range of higher positive case ratios and maintaining a very good baseline cost. Higher pooling factors present better economic outcomes only in the screening population with a relatively low prevalence of infection (<2.5% of positive cases in the population). It should also be taken under consideration that the cost of pooled testing is highly sensitive to the false positive ratio of the screening test; therefore, such a test must be highly sensitive for screening purposes but also provide high specificity.

**Figure 4.**
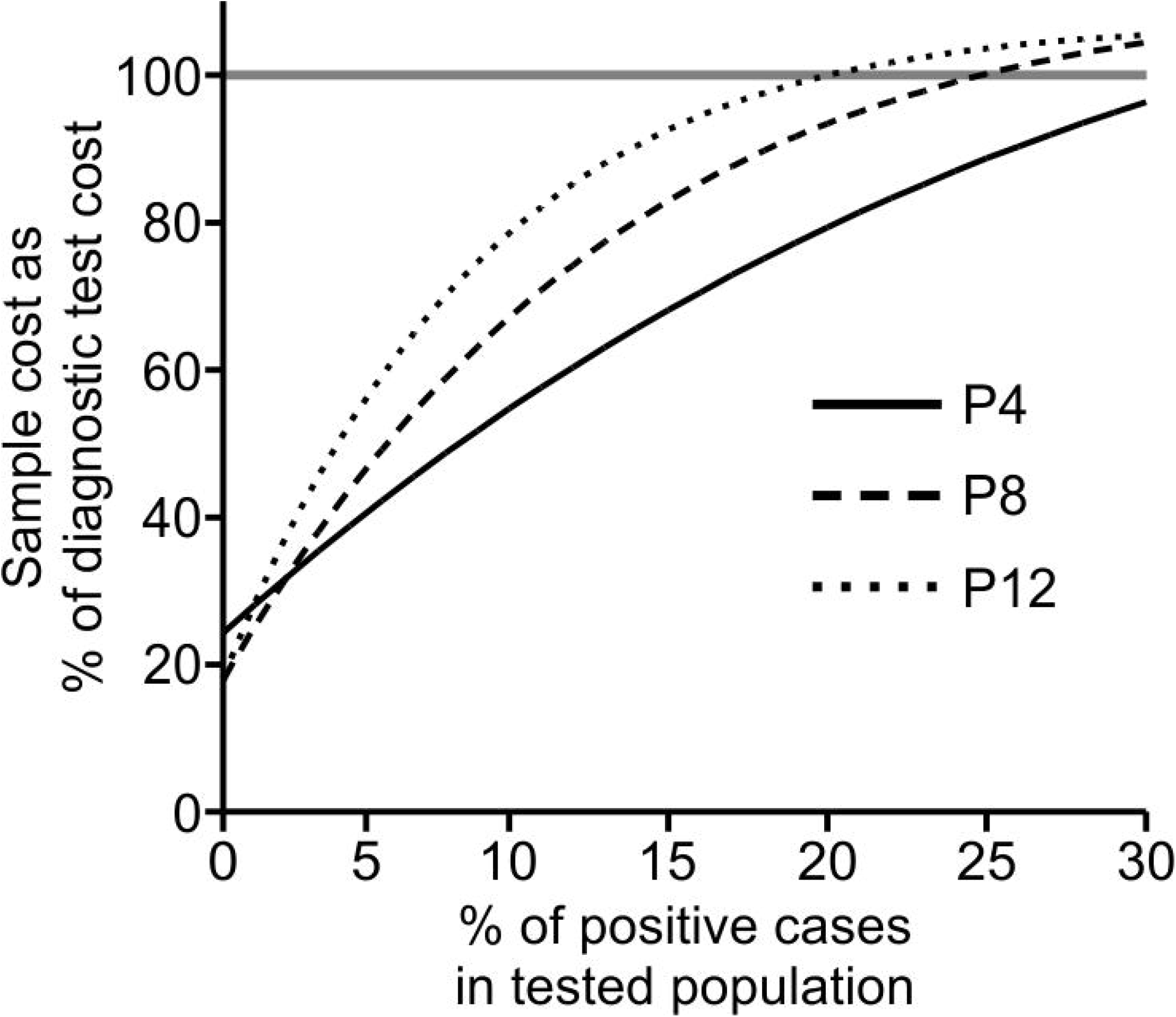
Economics of screening test. Comparative costs of testing using epidemic pooling tests and individual diagnostic tests employing a one-step, multiplex RT-qPCR SARS-CoV-2 commercial kit. Data are normalized against the cost of individual testing using commercial diagnostic tests (100%, horizontal line) and for both methods include the cost of reagents, disposable, PPE, labor, laboratory equipment and overheads.

### Validation of screening test in routine use

We performed multiple rounds of monitoring of employees of two institutions involved in the study for SARS-CoV-2 infection (involving a total of 113 people who decided to take part in our study). Testing was performed on weekly basis. All positive pools were unraveled, and individual samples were tested using a reference test (IVD certified commercial diagnostic test). In selected rounds, negative pools were also unraveled, and individual samples were tested with a reference test. The data on the performance of the screening test under such a scenario are presented in Figure 5. The screening test showed a 0.80% false positive rate, defined as the number of pools assigned as positive by the screening test outcome that did not contain at least one individual component categorized as positive or inconclusive in the reference test after unraveling the pool, divided by the total number of pools tested in screening. In 22 unraveled negative pools, there was no false negative defined as a pool tested as negative with at least one individual component categorized as positive or inconclusive in the reference test.

**Figure 5.**
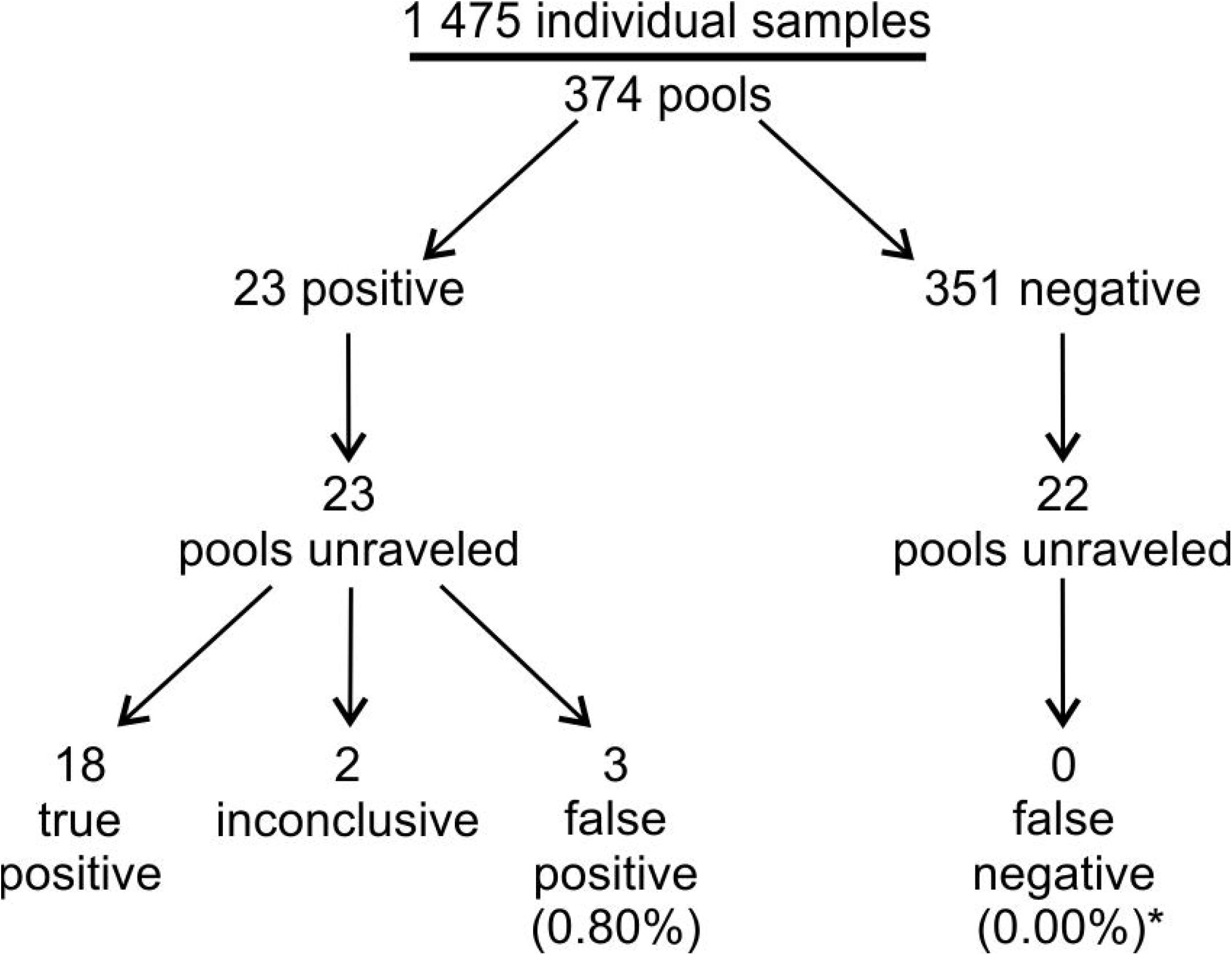
Results of routine use of the screening test; *false negative ratio calculated only from unraveled negative pools (which include 88 individual samples).

## Discussion

We have developed and characterized a qPCR-based method for the detection of SARS-CoV-2 dedicated to massive testing using pooled saliva samples. Tests have been employed in practice in a two-tier approach, in which each pool positive in the screening test is unraveled and individually tested using a standard diagnostic test. The idea of massive screening of populations for the presence of viruses is being explored in different countries with different testing methodologies. One potential method of such testing is based on simple point-of-care rapid tests such as antigen tests^7,8^ or LAMP-based genetic tests^9,10^. Such an approach has certain obvious advantages, such as decentralization of testing, fast turnover of test results, and relatively low cost. On the other hand, available, rapid individual tests do not have sufficient sensitivity to identify infected individuals at the earliest phase of infection (prior to onset of symptoms) and asymptomatic carriers^11^. Massive screening of populations performed with the use of such tests has resulted in unsatisfactory outcomes in several countries, among which testing of almost the entire country population of Slovakia is a well-publicized example^12^. On the other hand, testing over 7 million people in the Chinese city Qingdao in October 2020 that employed PCR-based, pooled, screening test allowed to nip the epidemics in the bud^13^. Comparison of the further development of epidemics in Slovakia and Quindao after the publication date of the articles cited above seems to further support the better outcome of Quindao screening (John Hopkins University SARS-CoV-2 worldspread map: https://coronavirus.jhu.edu/map.html, access April 07, 2021).

Our approach for massive PCR-based screening for SARS-CoV-2 is based on the detection of viral RNA in saliva. While nasopharyngeal swabs are still considered by the WHO as the diagnostic gold standard, there is already a bulk of literature data validating saliva as a diagnostic specimen. Several clinical studies comparing nasopharyngeal swabs and saliva as diagnostic specimens have been performed on a relatively large number of patients in different countries and have demonstrated comparable sensitivity and specificity of both diagnostic approaches^14,15,16,17,18^. This conclusion is also supported by a systematic review of 28 original reports^19^ and by a meta-analysis of clinical data representing 16 clinical trials performed on 5922 patients who showed 83.2% sensitivity of saliva-based tests compared to 84.8% nasopharyngeal swabs^20^. Taking into account comparable sensitivity, it should be stressed that saliva-based testing has a significant advantage over swabs in screening children in schools, where the discomfort of performing swabs may prevent compliance with routine testing^21,22,23^. It is also preferable in terms of mitigating the risk of exposure of medical personnel to infection, as self-collected samples could be directly delivered to designated drop-off points, which allow scattered sample collection without involving additional healthcare workers.

The proposed seminested qPCR with SYBR green-based detection of viral RNA based on the combination of the Ct threshold and range of Tm (Figure 2A and 2B) showed high sensitivity and specificity. The sensitivity allows for the detection of 5 copies of the viral genome (probability > 50%) and 95% probability of detection of 18.8 copies of the genome in 10 µl of isolated RNA, which is comparable to the sensitivity of qPCR detection based on fluorescent hybridization probes. According to the manufacturer manuals of both IVD reference tests used in this study (DiaPlexQ™ (SolGent) and Triplex (Vazyme)), both tests have a limit of detection of 200 genomes per milliliter of sample prior to RNA isolation (2 copies/10 µl) in nasopharyngeal swabs. Screening test in saliva showed the threshold of detection of 27 copies/10 µl. The direct comparison of published sensitivity thresholds is challenging because of different RNA isolation methods and different diagnostic specimens employed. Additionally, the limit of detection values published for a group of diagnostic assays officially approved until half of 2020 varied by 10,000-fold^24^. However, in side-by-side comparison, our test in saliva-based testing performs as well as Triplex (data not shown) and slightly better than DiaPlexQ™. Our data suggest that practical test sensitivity will be limited by isolation of the RNA step; samples with a lower number of viral particles may limit detection even to 5% of the sensitivity observed with the reference RNA solution and allow the detection of approximately 27 viral particles in 10 µl of saliva (>50% chance of detection) (Figure 3C). Relatively high sensitivity is matched by high specificity demonstrated *in silico* (Figure 1) and experimentally (Figure 3D). This method also showed good linearity within a wide range of viral RNA concentrations (Figure 3A) and viral load (Figure 3C). This is expected because SYBR green-based qPCR is well fitted for quantification of nucleic acids and makes the proposed method applicable in a quantitative approach for detecting the viral load in SARS-CoV-2-infected patients. In particular, quantitative measurements of SARS-CoV-2 RNA concentration in saliva are well correlated with the clinical outcome of infection^23,25^. High sensitivity of detection of viral RNA allowed for effective test application to pooled biological samples in this method, which demonstrated 100% specificity and 89.8% sensitivity in pools of saliva factored by 4, 6, and 8. This is consistent with published data on the application of fluorescent hybridization probe-based qPCR tests to pooled saliva samples, where there was 90-94% conformity with individual testing^26^. The test has already been implemented on a small scale for the routine testing of a group of 113 employees with promising results as the first tier of a two-tier testing approach. With such an approach, testing polled saliva samples shows significant cost savings compared to individual testing based on commercially available IVD RT-qPCR tests (Figure 4). Our results revealed that pooling saliva samples for the detection of SARS-CoV-2 infection support testing with substantial cost savings, especially at lower prevalence levels. The similar results have been obtained by other authors^18,27,28^. Implementation of the developed test for routine testing generated data for 1475 individual samples combined into 374 pooled samples that demonstrated very good test performance, with 0.8% false positives and undetectable false negatives for 88 individually analyzed samples (Figure 5). The lack of false negatives suggests that our pooled saliva-based qPCR testing may have a much lower false negative ratio than previously prognosed for saliva pools factored by 2-, 4-, 8-, 16 and 32 at the level of 10%^29^. It is worth mentioning that routine testing based on this test several times led to the detection of SARS-CoV-2 infection in asymptomatic persons, preventing them from coming to work and most likely decreasing the risk of virus transmission among coworkers. This difficult to objectively control but repeatable observation is well aligned with the output of an *in silico* model of epidemics in hospitals predicting that weekly screening by saliva-based PCR tests would be able to detect 95% of symptomatic and 30% of asymptomatic SARS-CoV-2 infections and reduce numbers of new infections^30,31^. Furthermore, the cumulative percentage of employees identified in prophylactic screening as SARS-CoV-2-positive was 15.9%, which is nearly threefold larger than the 6.21% of cumulative positive cases for the whole Polish population as of April 1, 2021 (John Hopkins University Map https://coronavirus.jhu.edu/map.html) and for a particular region (Voivodeship of Lodz with 6.53% of cumulative positive cases vs. 6.95% of the mean for the whole country, according to data from the Polish Ministry of Health at April 15, 2021). At the same time, the average percentage of positives per number of individual tests was 1.22%, which is well below the 5% maximum threshold suggested by the WHO as a prerequisite for effective monitoring of epidemic dynamics (https://www.who.int/docs/default-source/searo/indonesia/covid19/who-situation-report-11.pdf?sfvrsn=3e0fb6c8_2, access April 16 2021). This is once again in contrast with the percentage of positives reported in diagnostic tests in Poland that, since the middle of October 2020, continuously exceeded 20% with a peak of 59% at 16.11.2020, and a temporary fading period, with a positive level of approximately 10-15% at the turn of January and February of 2021 (Polish Ministry of Health). Taken together, these data support the validity of the proposed analytical approach for prophylactic screening of selected populations for infection during COVID-19 epidemics.

## Data Availability

The authors confirm that the summarized data supporting the findings of this study are available within the article. The raw data are available from the corresponding author, Jarosław Dastych, upon reasonable request.

## Acknowledgements

We acknowledge Prof. Krzysztof Pyrć from Jagiellonian University for sharing standardized SARS-CoV-2 RNA sample. The following reagent was deposited by the Centers for Disease Control and Prevention and obtained through BEI Resources, NIAID, NIH: SARS-Related Coronavirus 2, Isolate USA-WA1/2020, Heat Inactivated, NR-52286. The following reagents was obtained through BEI Resources, NIAID, NIH: Genomic RNA from Human Respiratory Syncytial Virus, A2001/3-12, NR-44227; Genomic RNA from Human Respiratory Syncytial Virus, B1 BPR-348-00, NR-4052; Genomic RNA from Human Coronavirus NL63, NR-470; Genomic RNA from Human Coronavirus 229E, NR-52726.

## Abbreviations

RT: reverse transcriptase
RT-qPCR: quantitative reverse transcription polymerase chain reaction
IVD: *in vitro* diagnostics (certificate)
dNTP: deoxyribonucleotide triphosphate
DEPC: diethyl pyrocarbonate
GAPDH: glyceraldehyde 3-phosphate dehydrogenase
Tm: melting temperature
Ct: threshold cycle (in quantitative PCR)
CI: confidence interval
HBSS: Hanks’ balanced salt solution
PPE: personal protective equipment

